# Quantifying Effects, Forecasting Releases, and Herd Immunity of the Covid-19 Epidemic in S. Paulo – Brazil

**DOI:** 10.1101/2020.05.20.20107912

**Authors:** S. Celaschi

## Abstract

A simple and well known epidemiological deterministic model was selected to estimate the main results for the basic dynamics of the Covid-19 epidemic breakout in the city of São Paulo – Brazil. The methodology employed the SEIR Model to characterize the epidemics outbreak and future outcomes. A time-dependent incidence weight on the SEIR reproductive basic number accounts for local Mitigation Policies (MP). The insights gained from analysis of these successful interventions were used to quantify shifts and reductions on active cases, casualties, and estimatives on required medical facilities (ITU). This knowledge can be applied to other Brazilian areas. The analysis was applied to forecast the consequences of releasing the MP over specific periods of time. Herd Immunity (HI) analysis allowed estimating how far we are from reaching the HI threshold value, and the price to be paid.

## Introduction

This work aims to shed some extra light in understanding the dynamics of the COVID-19 epidemics in Brazil, particularly in the city area of São Paulo with its 12.2 million inhabitants. The first patient in Brazil was tested positive in São Paulo who had returned from Italy. Since then, were officially confirmed 233.142 cases, and 15,644 deaths in Brazil (May, 16 2020), and 61,183 cases, and 4,688 deaths in S. Paulo. The public response to the pandemic has been the introduction of mitigation policies to ensure quarantine social distancing, such as closing schools, restricting commerce, and home office. No country knows the true number of people infected with COVID-19. All is known is the infection status of those who have been tested. The total number of people that have tested positive – the number of confirmed cases – is not the total number of people who have been infected. The true number of people infected with COVID-19 is much higher. The number of those who have been tested positive (by May 1st) in Brazil, 0.46/thousand, is very low compared to other countries [1] (Fig. 1).

**Figure 1.**
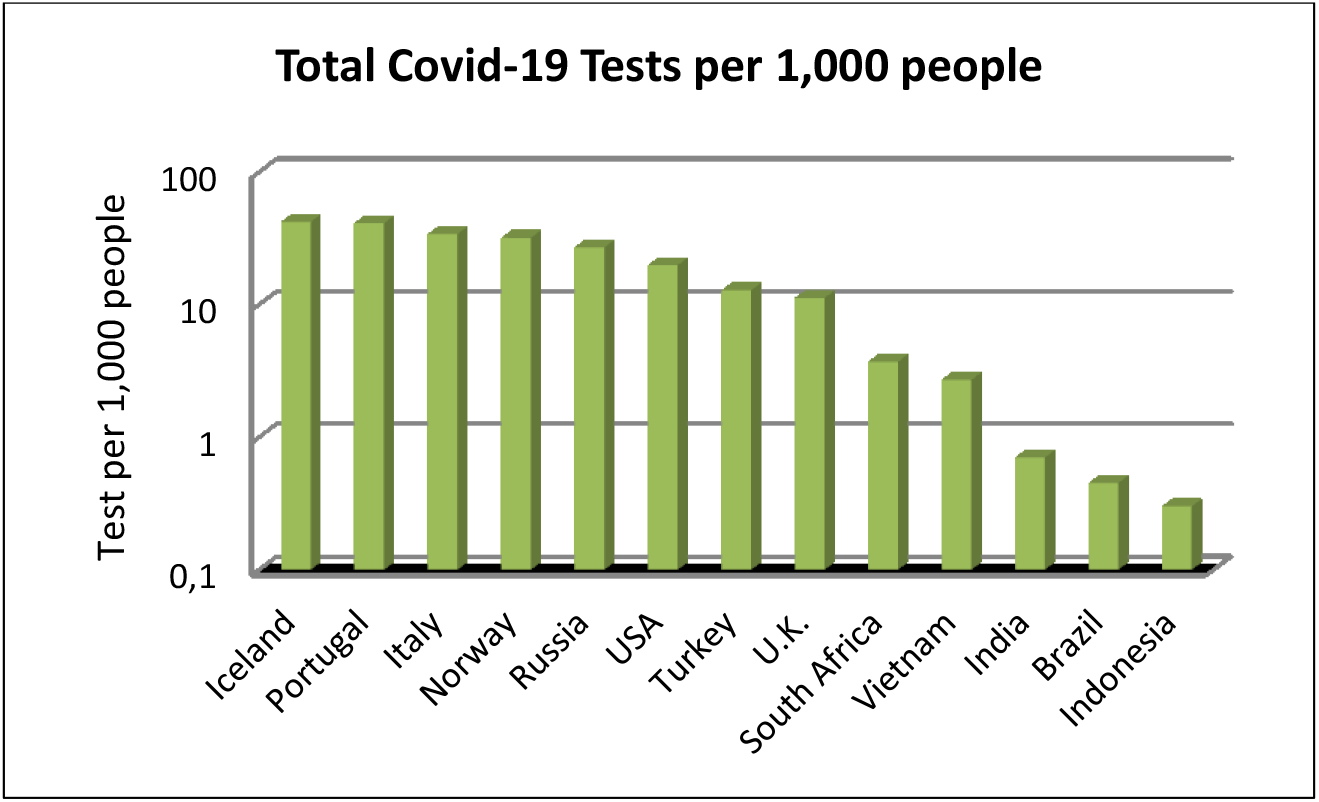
*Data (relative to May, 1^st^) collected from www.ourworldindata.org [1]*.

The response to a new pandemic, such as Covid-19, can be based on four major actions: 1) surveillance and detection; 2) clinical management of cases; 3) prevention of the spread in the community; and 4) maintaining essential services. Actions across the four pillars complement and support one another. In principle if the virus is left to infect people without any containment measure, the population may acquire immunity in one semester or less. However, hospital intensive care services would lack the capacity to deal with the sudden, large inflow of severely ill people resulting in a very large number of deaths. Mitigation strategies aim to slow the disease, and to reduce the peak in health care demand. This includes policy actions such as social distancing, lock-down determination, and improved personal and environmental hygiene.

Studies as this presented here, consistently conclude that packages of containment and mitigation measures are now days an effective approach to reduce the impact of the Covid-19 epidemic. Epidemiological models are commonly stochastic, diffusive-spatial, network based, with heterogeneous sub-populations (meta-population approaches) [2, 3, 4]. However, the parameters of dynamical and deterministic models, such as SIR and SEIR, are more directly related to and interpretable as physical processes [5, 6]. On the other hand, deterministic models impose restrictive analysis, once the dynamics of the host population and the virus are not deterministic. The population has free will, and the virus undergoes “random” mutations.

The intent of this work was to build a simple epidemiological tool to estimate the main results for the basic dynamics of the Covid-19 epidemic breakout. The methodology employed is the application of the deterministic and discrete SEIR Model to characterize the Covid-19 outbreak in São Paulo – Brazil. The model accounts for the following 5 groups: Susceptible, Exposed, Symptomatic infected, Asymptomatic infected, and Removed or recovered. A time-dependent incidence weight on the SEIR basic reproductive number R(t) was used to account for dynamical transmission behavior to model and quantify local Mitigation Policies (MP). The insights gained from analysis of these successful interventions can be used to predict results for the MP of other Brazilian regions.

Recent published data [7, 8] from January, 1st, 2020 to May 8, 2020 was used to adjust all the model parameters aiming to forecast the evolution of the COVID-19 pandemic in the city of Sao Paulo - Brazil. The model provides predictions of the time series of infected individuals and fatalities in the area studied. Simulations of midi-term scenarios of the epidemic outbreak were done dependent on the level of confinement policies. Forecasts show how confinement policy alters the pattern of contamination, and suggest the existence of post epidemic periods.

Application of mathematical models to disease surveillance data can be used to address both scientific hypotheses and disease-control policy questions [9]. Such models have been used to estimate the demand for hospital beds, ICU days, number of critical equipments, and the need and required extent of governmental intervention. A model is only as good as the assumptions put into it. Clearly, there are phenomena of the COVID-19 epidemic that are not yet understood. Models are constantly being updated and improved.

The SARS-CoV-2 is a membrane protected, single-stranded RNA virus. It is commonly referred to by the name of the disease it causes, which is COVID-19. The incubation period is defined as the time between infection and onset of symptoms. It is estimated as the time between exposure and report of noticeable symptoms. Currently, the incubation period for COVID-19 is somewhere between 2 to 5 days after exposure [13]. More than 97 percent of people who contract SARS-CoV-2 show symptoms within 12 days of exposure. For many people, COVID-19 symptoms start as mild symptoms and gradually get worse over a few days. Other large and unknown fraction of exposed people is asymptomatic. The story of the COVID-19 outbreak is ongoing. Our knowledge of this novel virus is in a state of flux. Every week seems to bring additional important medical and epidemiological information.

COVID-19 has a latent or incubation period, during which the individual is said to be infected but not infectious. Members of this population in this latent stage are labeled as Exposed (but not infectious) here on. The model with the Susceptible, Exposed, Symptomatic, Asymptomatic, and Removed groups is our SEIR Model. During the initial 20 days exponential phase of growth, the SP data shows that in 2.3 ± 0.1 days the number of symptomatic infected people doubles. During this initial exponential period, the model confirms the predicted values for the incubation, and immunization periods of time.

## The SIR and our SEIRD Models

The SIR model [5] is one of the simplest compartmental models, and many other models are created from this basic formulation. The model consists of three compartments: S for the number of susceptible, I for the number of infectious or active cases, and R for the number of recovered, deceased (or immune) individuals. This model is reasonably predictive for infectious diseases that are transmitted from human to human. In epidemics or pandemic outbreaks, the numbers of susceptible, infected and recovered individuals varies with time (even if the total population size remains constant). For a specific disease in a specific population, these functions may be worked out in order to predict possible outbreaks and bring them under control. For many important viral infections, there is a significant incubation period during which individuals have been infected but are not yet infectious themselves. During this period the individual is in a new compartment E (for exposed) as in the SEIR model.

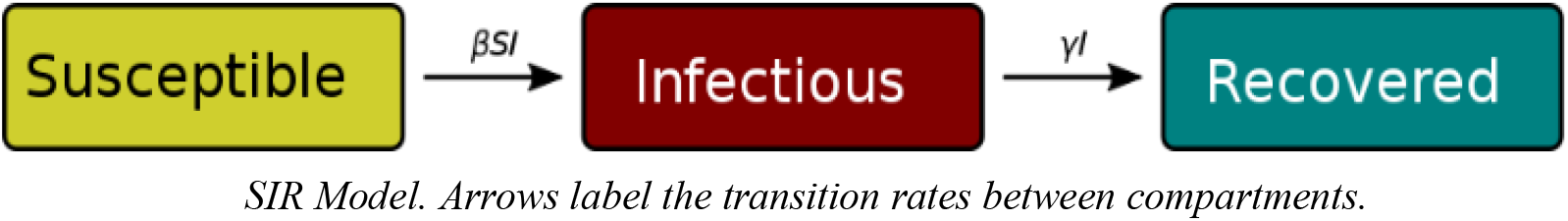

*SIR Model. Arrows label the transition rates between compartments*.

Our S-E-Ia-Is-R version of the SEIR model [10] describes the spread of a disease in a population split into five nonintersecting groups:

(S) Susceptible: The population that can be exposed to the disease;
(E) Exposed: Group in the latent or incubation stage exposed to virus but not infectious;
(I_s_) Symptomatic Infected: Group of individuals who are infected, and account to the official number reported cases;
(Ia) Asymptomatic Infected: Group of individuals who are infected, and do not account to the official number of cases reported;
(R) Removed: Group of individuals who are recovered from the disease.

Due to the evolution of the disease, the size of each of these groups change over time and the total population size N is the sum of these groups

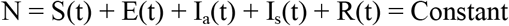

At the initial exponential outbreak, let *β_o_* be the average number of contacts (per unit time) multiplied by de probability of transmission from an infected person. Let *β(t) = β_a_(t) = β_s_(t)* be the infection rate that models temporal Mitigation Policies (MP). Let *ψ(t)* quantify the MP in terms of social distancing plus the use of personal protective equipment (PPE). Let *σ_o_* be the rate that exposed individuals get infected. Let *γ_a_* and *γ_s_* be the removed rates, which are the rates that infected individuals (symptomatic and asymptomatic) recover or die (only symptomatic), leaving the infected groups, at constant per capita probability per unit of time. Let *ξ_a_* being the fraction of asymptomatic individuals. Let *N* be the susceptible population considered in the study. Based on these definitions, we can write the model as:

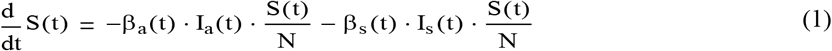

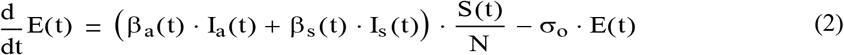

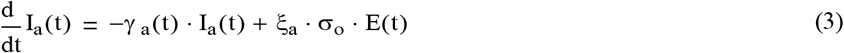

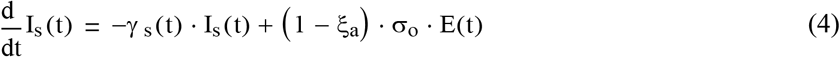

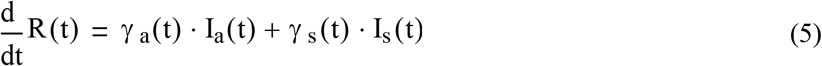

constrained to the following initial conditions: S(0) = N, E(0) > 0, I_a_(0) = 0, I_s_(0) = 0, R(0) = 0.

By the time this study was conducted, information on the number of asymptomatic infected individuals was unknown. So, some parameters could not be determined in order to fully apply our S-E-I_a__I_s__R version of the SEIR model. Later on, this restriction may be removed once the required information on Covid-19 asymptomatic hosts will be reported by testing an expressive fraction of the population. The following assumptions are established in order to continue the analysis: *β(t) = β_a_(t) = β_s_(t), γ = γ_a_ = γ_s_*, and 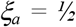, and total number of infected people will be *I(t) = (1-ξ_a_) I_s_(t) + ξ_a_I_a_(t)*. The set of ordinary differential equations (Eqn. 1 to 5) is reduced to the standard SEIRD model,

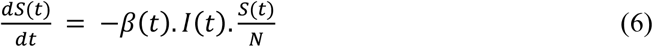

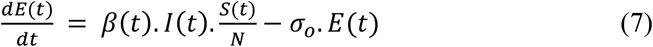

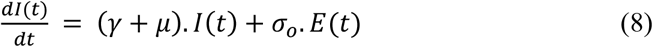

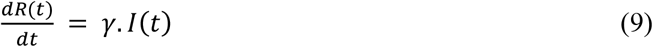

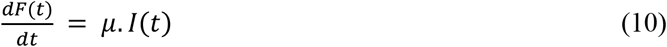

An extra equation (Eqn.10) has been added to account for fatalities to complete the SEIRD Model. As before, *S(t), E(t), I(t), R(t), and F(t)* are respectively daily numbers of susceptible, exposed, infected or active cases, removed or recovered individuals, and fatalities (deaths). *S(t)* + *E(t) I(t) R(t) + F(t) N Constant*. The constant *N* assumption is very restrictive, and limits model’s coverage. Releasing this assumption goes beyond the scope of this study. *R_N_(t) = β(t)/γ_o_. R_N_(t=0) = R_o_*, is defined as the Basic Reproductive Number. This number quantifies the expected number of new infections (these new infections are sometimes called secondary infections) that arise from a typical primary case in a completely susceptible population *N*, where all individuals are susceptible.

## Methodology

Official data, from March 1st to May 15, 2020, provided by the Ministry of Health of Brazil [7], and Sao Paulo government [8] was considered to estimate part of the epidemiological parameters that govern the dynamics established by Eqs. (6), (7), (8), and (9). By the lack of information about the asymptomatic individuals, the mortality rate in the model is evaluated over the symptomatic ones. All model parameters were estimated by minimizing the mean squared quadratic errors.

A key parameter in deterministic transmission models is the reproductive number *R_o_*, which is quantified by both, the pathogen and the particular population in which it circulates. Thus, a single pathogen, like the SARS-CoV-2, will have different R_o_ values depending on the characteristics and transmission dynamics of the population experiencing the outbreak. When infection is spreading through a population that may be partially immune, it has been suggested to use an effective reproductive number R, defined as the number of secondary infections from a typical primary case. Accurate estimation of the *R* value is crucial to plan and control an infection [11].

The methodology to estimate *R* follows: The exponential growth rate of the epidemic, *r* was obtained from the early stages of the epidemic in Sao Paulo, such that the effect of control measures discussed later will be relative to post stages of this outbreak. This assumption is implicit in many estimative of *R*. The growth rate *r = 0.31* ± *0.02* of infected people was estimated applying the Levenberg-Marquardt method [12], to data of symptomatic infected people (Fig. 2), during the first 15 days of exponential growth according to the expression *I(t) = Io.exp(-r.t)*.

**Figure 2.**
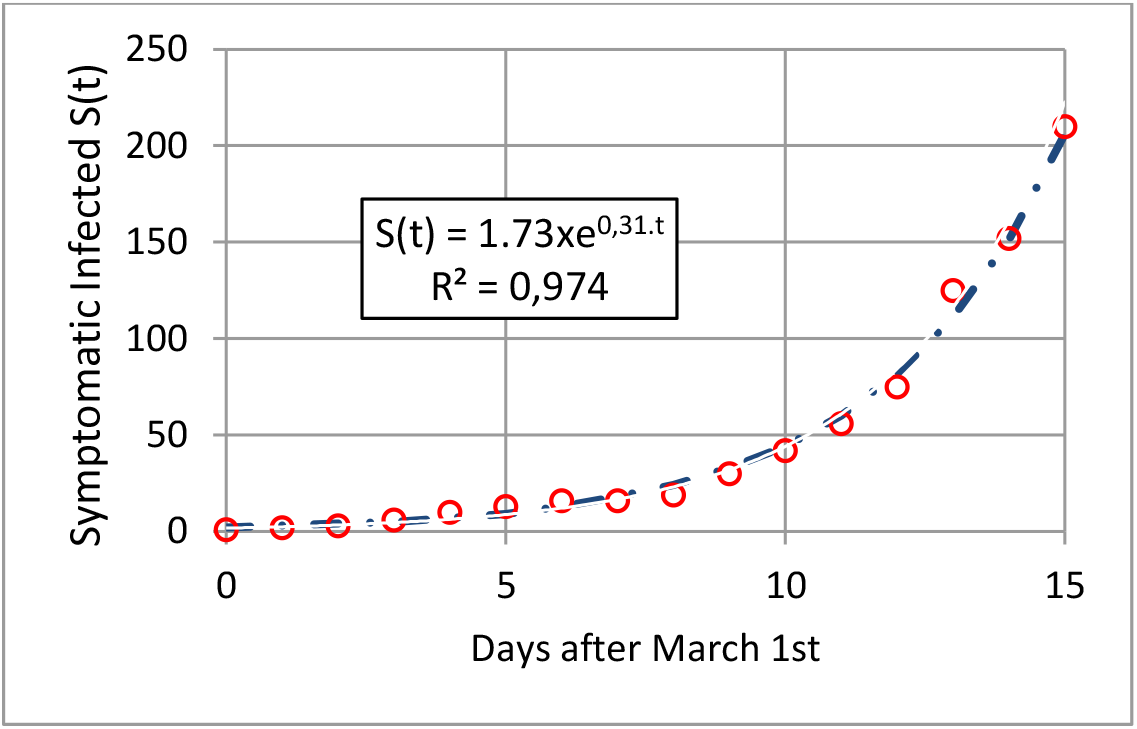
*Exponential fitting of the initial growth of symptomatic infect people. The growth rate r = 0.31 ± 0.02 was estimated applying the Levenberg-Marquardt method*.

The time period required to double the number of symptomatic cases is straightforward given by *ln(2)/r = 2.3* ± *0.1 days*. The basic Reproductive Number *R_o_* — *2.53* ± *0.09* ^vas esti^nated according to; “In an epidemic, driven by human-to-human transmission, whereas growing exponentially, in a deterministic manner, the incidence *I(t)* can be described by the Renewal Equation”, or the Lotka-Euler equation [13, 14]:

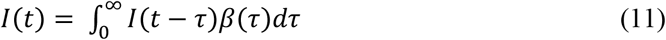

Where β(τ) is the mean rate at which an individual infects others a time after being infected itself. Substituting into Equation (11) an exponentially growing incidence, *I(t) = I_o_.exp(r.t), I_o_=1*, gives the condition,

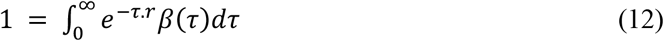

where

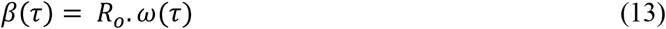

*ω(τ)* is the generation time distribution, i.e. the probability density function for the time between an individual becoming infected and their subsequent onward transmission events. *R_o_* is the basic reproduction number. If the exponential growth rate *r* and the generation time distribution *ω(τ)* have been estimated, *R_o_* is readily determined from Eqn. (12), as

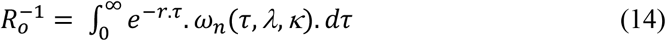

The term that appears in the right-hand side of this equation is the Laplace transform of the integrand function. More specifically, it is known as the Momentum Generating Function of this distribution. As in [13], a normalized Wiebull generation distribution is adopted (Eqns. 15, 16 and 17), with mean = 0.89, median = 3.1, and peak value at 2.3 days (Fig. 3).

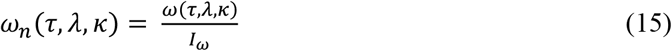

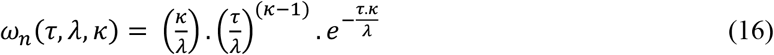

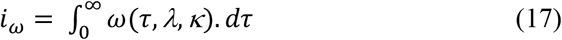

**Figure 3.**
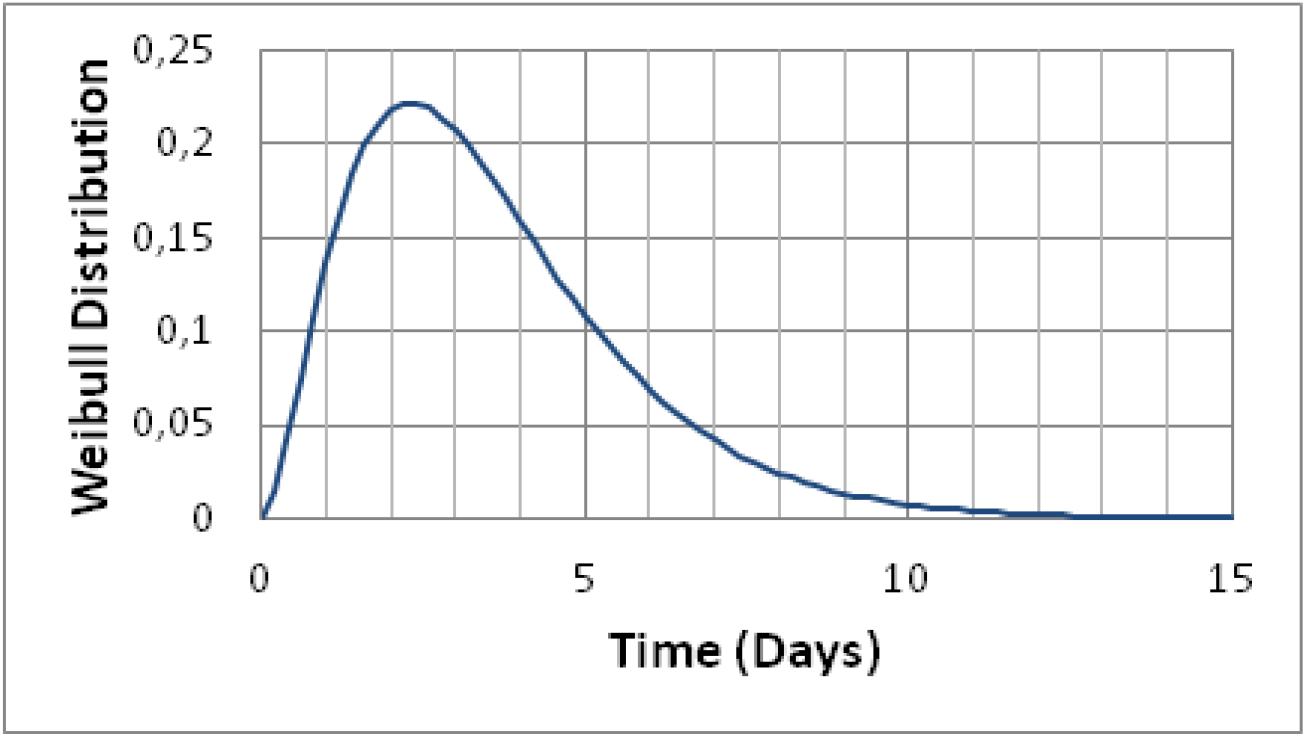
*Distribution of generation times. Our data was described by the Weibull distribution with mean = 0.89, median = 3.1 days, and peak value 2.3 days. Dates of symptom onset with intervals of exposure for both source and recipient (when available) were collected in [13] in order to select the best distribution*.

In short, the values of the parameters governing SEIRD model are: *β(t), γ, σ_o_, μ, and N*. The SEIRD dynamics is constrained to the following initial conditions *S(0) = N, E(0) > 0, I(0) = 0, R(0) = 0*. March 1^st^ 2020 was considered the first day (day zero) to model the epidemic outbreak at the city of Sao Paulo. As proposed by Bastos et al. [10], the temporal impact of the confinement policy was considered weighting the initial transmission factor *β_o_* by *ψ(t)* fitted to data, and adjustable to allow releases of the MP (Fig. 4a) [7,8]. This leads to β factor as a temporal function *β(ψ_o_, t_o_*, *t_1_, t_2_, φ, t)*, where *ψ_o_, t_o_, t_1_, t_2_, and φ* are values set by the MPs considered in this report.

**Figure 4.**
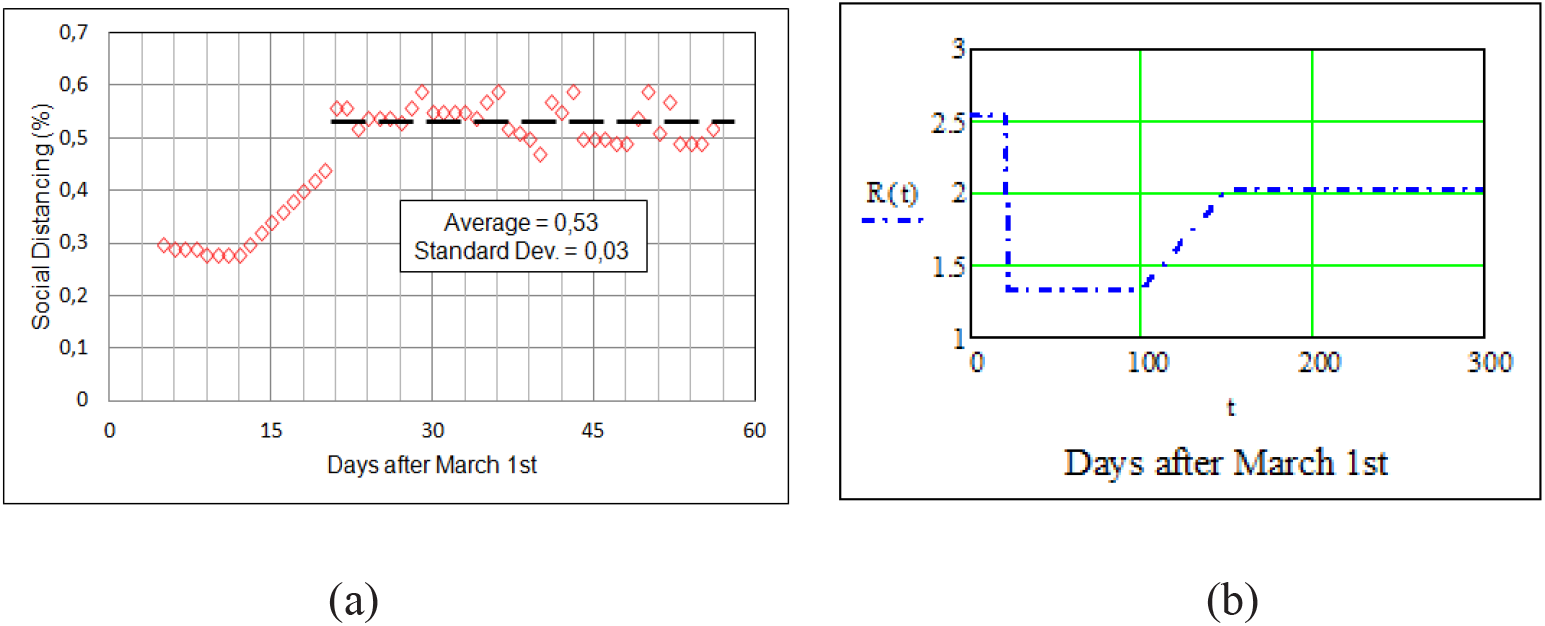
*Quantification of the MP application on the city of S. Paulo by Social Distancing (SD) measures (a), Reproductive Basic Number R(t) modeling the Social Distancing effective change after March 22^nd^ (b). Blue point and dashed line represent the progressive return to R_o_ after day 100*.

Accordingly, *R(t)=ψ(t).R_o_* becomes dependent on the confinement policy (Fig. 4b). The effectiveness of this policy may be quantified by a social distancing factor defined as *SD = 1 − ψ(t)*. Furthermore, by lack of information about asymptomatic hosts, a value of 50% of the exposed hosts was assumed.

## Discussion and Forecasts

The fittings of data on symptomatic infected individuals, and fatalities to the SEIRD model are show in Fig. 5. The fitting values are: *R_o_ = 2.53, β_o_ = 0.9, γ_o_ = 0.339, ψ_o_ = 0.525, μ = 0.017, σ_o_ = 0.5, and t_1_ = 22*. The fitting to the infected individuals (logarithmic scale) presents standard deviation SD = 0.05 and root mean square RMS = 0.07. The exponential rate of incubation *1/σ_o_* was assumed here as 2 days. Important to mention that the model was also applied assuming *1/σ_o_= 4* days [13], and fixed values for *R_o_ = 2.53, ψ_o_ = 0.525, μ = 0.017, σ_o_ = 0.5, and t_1_ = 22*. The new fittings to data, preserving *R_o_ (β_o_ = 2.445, and γ = 0.923)*, led essentially to the same results.

Figure 5 shows reductions in exponential growths of infected and death rates after the third week. This reduction clarifies the effectiveness of the confinement policy, when social distancing took place. In fact, after day 22, the data shows reduction in the tax of transmission. So, was defined as the initial date to estimate the parameter ψ_o_, keeping all model parameters as previously estimated [10]. The transmission rate *β(t)* is reduced to approximately 52% of its original value *β_o_*, leading to a MP of 48% in accordance to the data shown in Figure 4. Final values for asymptomatic infected individuals and fatalities, at day 190, are respectively 110,000, and 10,500. Taking into consideration all the hosts, the average rate of lethality is 4.5 ± 0.5 %, in agreement to recent published data [14].

**Figure 5.**
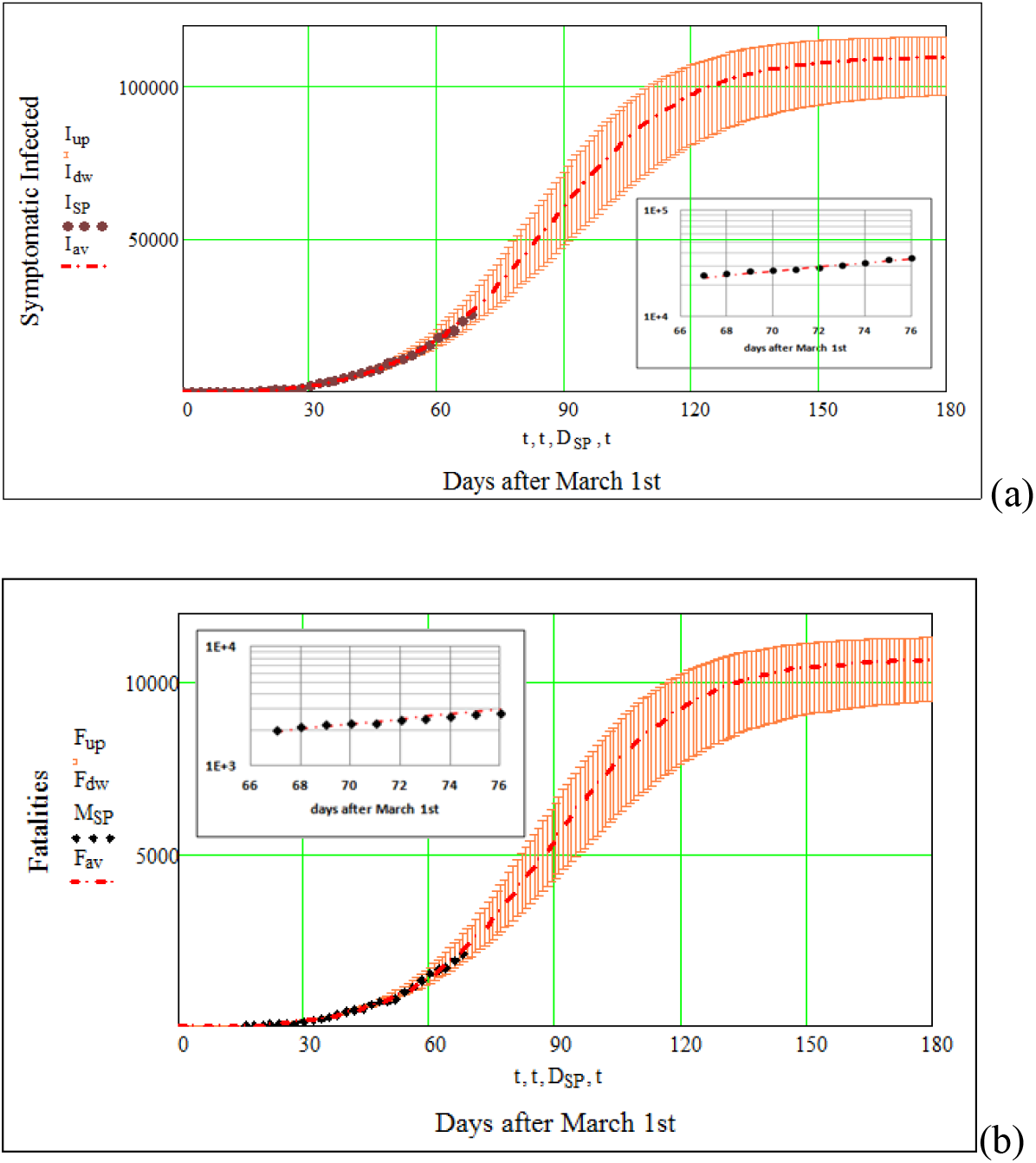
*Official data on symptomatic infected individuals (a), and fatalities (b) are shown by black circles, and diamonds respectively. Error bars account for the RMS values on the SEIRD parameters. The fitting values to SEIRD model shown by red dashed line and error bars are: R_o_ = 2.53* ± *0.09, β_0_ = 0.913* ± *0.018, γ= 0.316± 0.01, μ = 0.017, ψ_0_ = 0.525* ± *0.03, σ_o_ = 0.50* ± *0.02, and t_SD_ = 22 days. The mean fitting to the infected individuals (logarithmic scale) presents standard deviation SD =0.05 and root mean square RMS = 0.07. <u>Data points for the last 10 days (inserts) were added after the model was complete, granting confidence on outcomes.</u>*

The major results obtained for the epidemic of COVID-19 in the city of S. Paulo, maintaining the MP for the entire period, are: At least 4% of susceptible persons; R_o_ = 2.5, the average number of symptomatic people infected by a sick person; Projected lethality rate ~ 5 %; Projected symptomatic infected hosts 110,000: Projected fatalities 9,800, keeping 48% SD constant.

Figure 6 presents data on the daily number of new symptomatic infected, compared to as predicted from modeling. The large data scattering on day to day-plus differences comes from the available reported cases. The number of Intensive Care Units (ICU) was estimated to be proportional to the weekly number of symptomatic infected. The proportionality constant in Eqn. (18) was determined as the ratio of occupied ICU beds (SARG - COVID-19) to the number of new infected during the 17^th^ week, where SARG = 8469 [7].

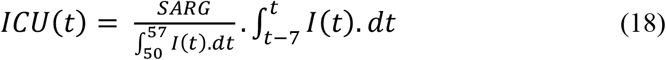

**Figure 6.**
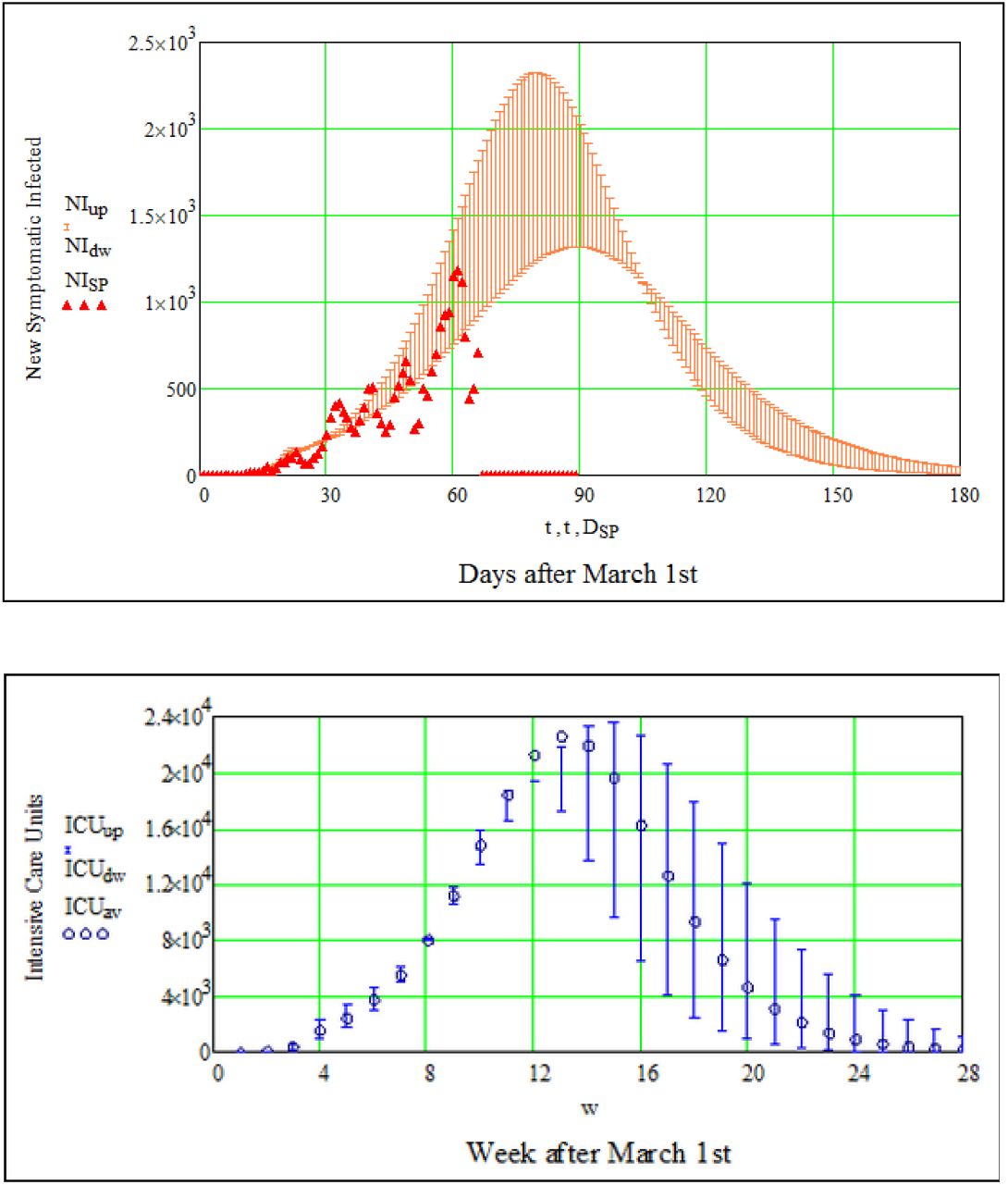
*Data on the daily number of new symptomatic infected compared to predictions of the SEIRD model (a). The large data scattering on day to day-plus differences comes from the available reported cases. The number of Intensive Care Units (ICU) was estimated to be proportional to the weekly number of symptomatic infected (b). Permanence of SD after day 22^nd^ was assumed. Error bars account for the RMS values on the SEIRD parameters*.

Prediction on the average weekly number of intensive care units (ICU) is also included in Fig. 6b by open blue circles. Error bars account for the RMS values on the SEIRD parameters. The maximum weekly number of ICU beds for COVID-19 is predicted to be approximately 24,000 by the first week of June, 2020.

Regarding fatalities, average data on daily new casualties is compare to model prediction on Fig.7a. Do to large data scattering; a 5 days weight moving average was applied to data on Fig. 7a. Keeping the MP, as modeled after March 22^nd^, the maximum number of new daily fatalities is predicted to happen by the last week of May 2020. To compare this outbreak regarding other countries, we present in Fig 7b the number of fatalities per thousand inhabitants recently published [16].

**Figure 7.**
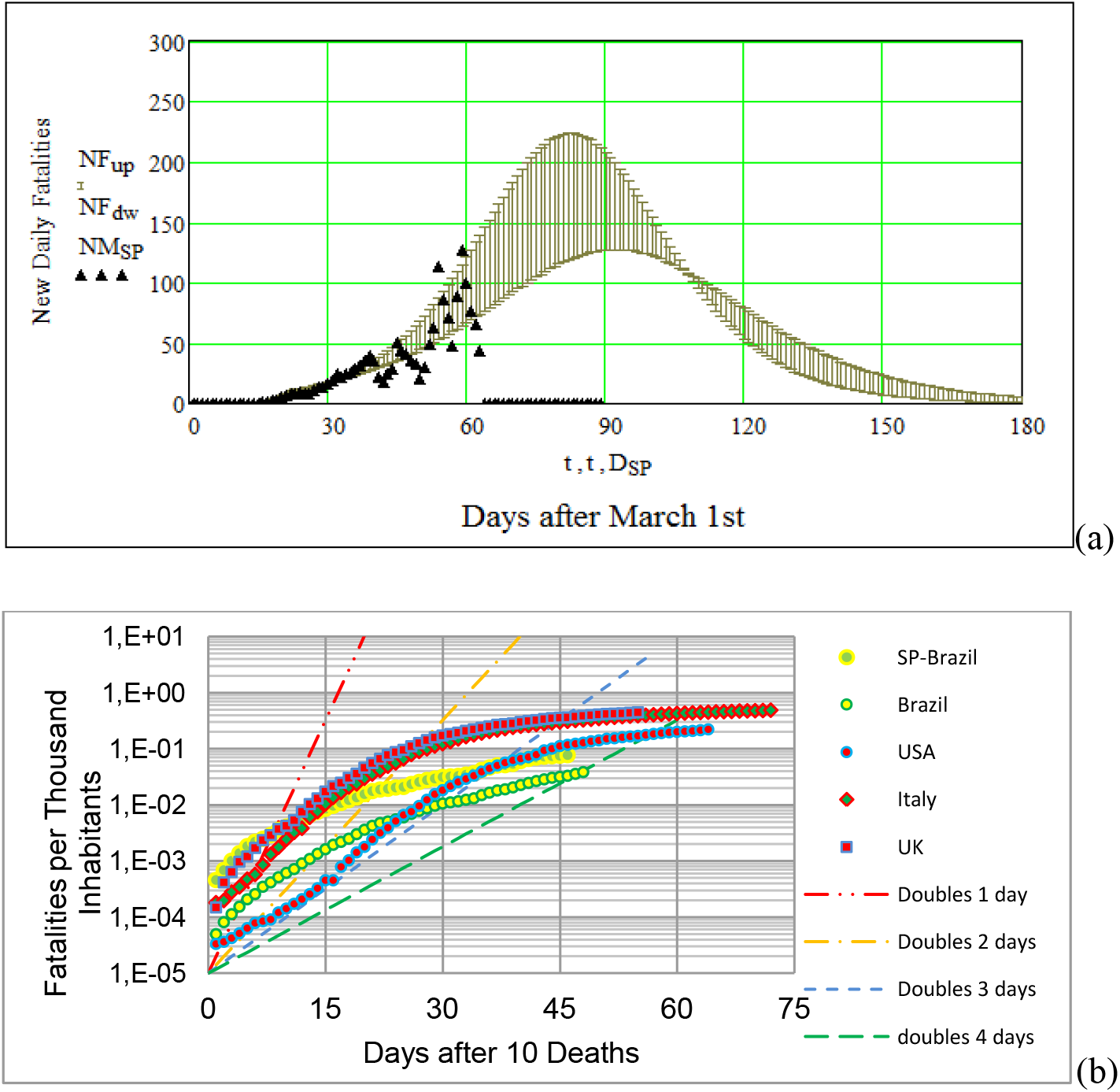
*(a) Data on daily new fatalities is compare to model prediction. Keeping the MP as modeled after March 22^nd^. The RMS values to SEIRD model are shown by black error bars The maximum number of new daily fatalities is predicted to happen by the last week of May 2020. (b) Number of fatalities per thousand inhabitants reported from other countries. The fatality raise in the city of S. Paulo is shown for comparison*.

The number of susceptible individuals *N = 500,000*, which represents 4.1% the population of Sao Paulo city, was considered to be the minimum number to fit the available data on new symptomatic hosts. As early mentioned, the constant *N* assumption restricts our analysis, and forecasts. Since the number of sub notifications is a reality well accepted, we present in Table 1 suggested results folding *N* by a factor of three.

**Table 1.** *Folding N by a factor of 3, peaks the active cases2.5 weeks later, and raises the number of immune hosts, ITU units, and fatalities by the same factor*.

| % Population | # Immunes<br>($\times 10^5$) | Peak Infection<br>(day) | Max # ICU<br>($\times 10^4$) | # Fatalities<br>($\times 10^4$) |
| --- | --- | --- | --- | --- |
| 4.1 | 2.1 | 87 | 2.2 | 1.0 |
| 6.2 | 3.1 | 95 | 2.8 | 1.5 |
| 8.2 | 4.2 | 100 | 3.6 | 1.9 |
| 10.3 | 5.2 | 103 | 4.4 | 2.4 |
| 12.3 | 6.2 | 107 | 5.2 | 2.9 |

In order to quantify the MP imposed at the city of Sao Paulo, a sequence of 3 plots presented by Fig. 8 demonstrate the effects of SD progressive releases imposed by state regulations. The sequence shows the daily numbers of additional symptomatic infected, and deaths. Social distancing of 48% was set as a constant from day - 22^nd^ on. Selected values for beginning a linear progressive SD release (t_2_), ending it (t_end_) to a final value of 24% are shown in Figs. 8a, 8b, and 8c. The sequence forecasts additional daily numbers of symptomatic infected (red line), and additional deaths (black line). The prospected accumulated additional fatalities for t_2_ = {90, 100, 110}, and t_end_ = t_2_ + 30 days are respectively 48, 36, and 32%.

**Figure 8.**
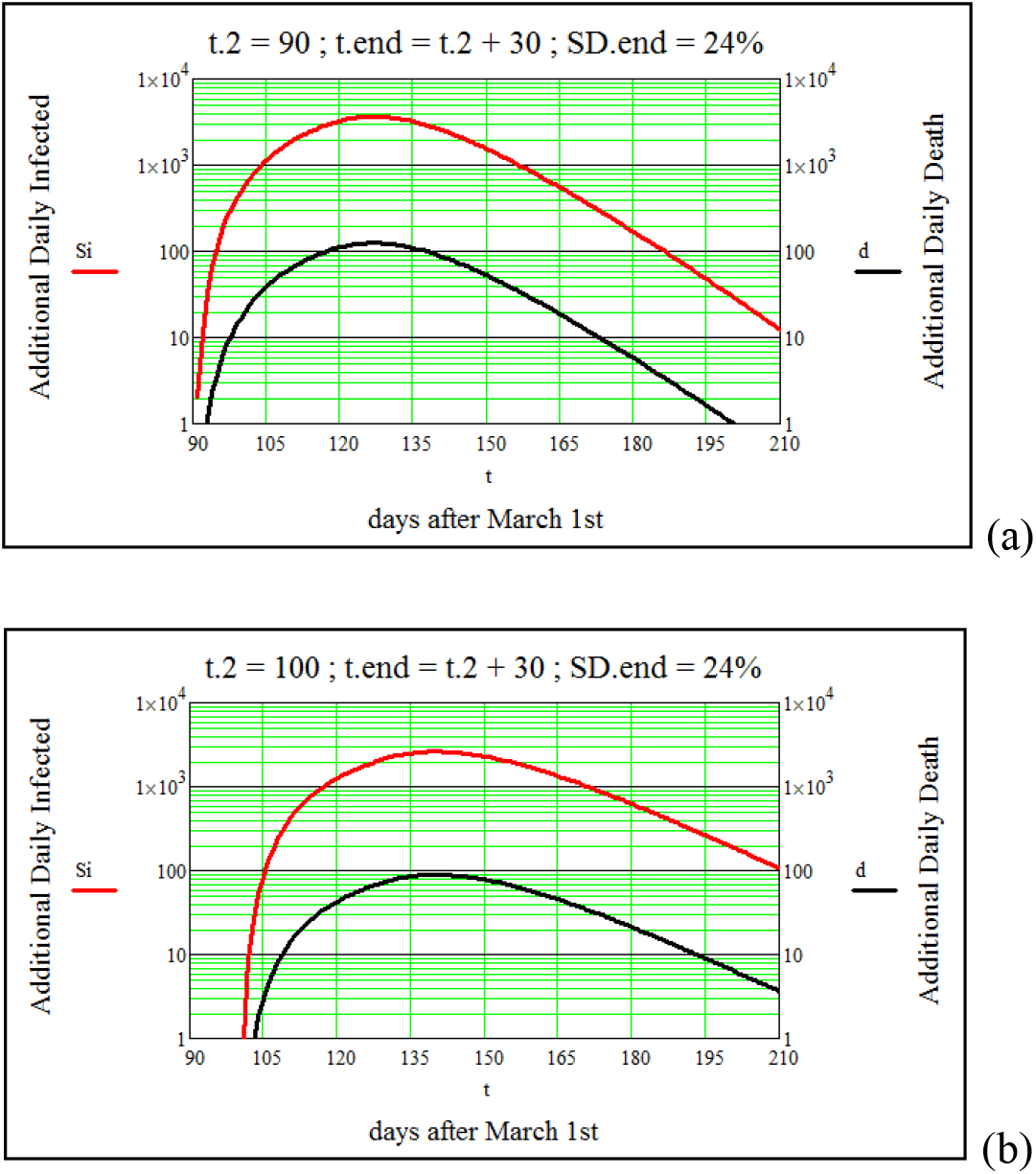
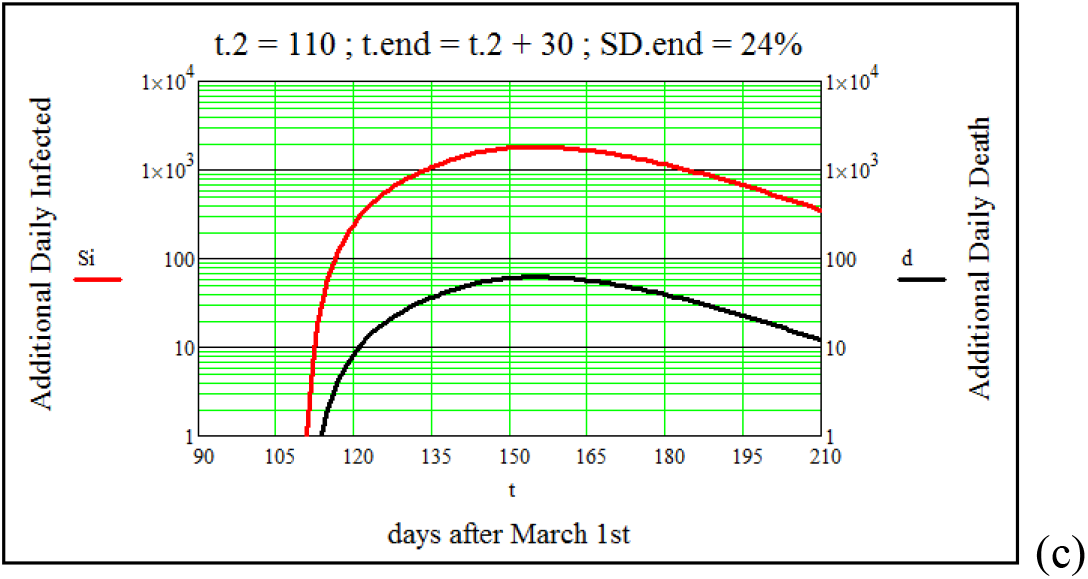
*The sequence of figures demonstrates effects of releasing the 48% social distancing set constant from day - 22^nd^. Selected values: t_2_ - beginning progressive SD release; t_end_ - end SD release, final SD = 24%. The sequence forecasts additional daily numbers of symptomatic infected (red line), and additional deaths (black line)*.

The analysis can also be applied to forecast the consequences of releasing the MP over a longer period of time. Figure 9 illustrates this simulation by a linear and progressive MP release starting by the end of the third month, and ending ten months later, when the reproductive number returns to its initial value R_o_. Surprisingly or not, the model suggests an “endemic” outbreak of Covid-19 as shown in Fig. 9 by the presence of the second peak ~11 months after the first one. The number of infected individuals is estimated to be 55% lower compared to the first outbreak. This is a result of a partial reduction of 58% on the initial number of susceptible individuals. As recently published by US National Library of Medicine, National Institutes of Health, agencies worldwide prepare for the seemingly inevitability regarding the COVID-19, to become endemic [15].

**Figure 9.**
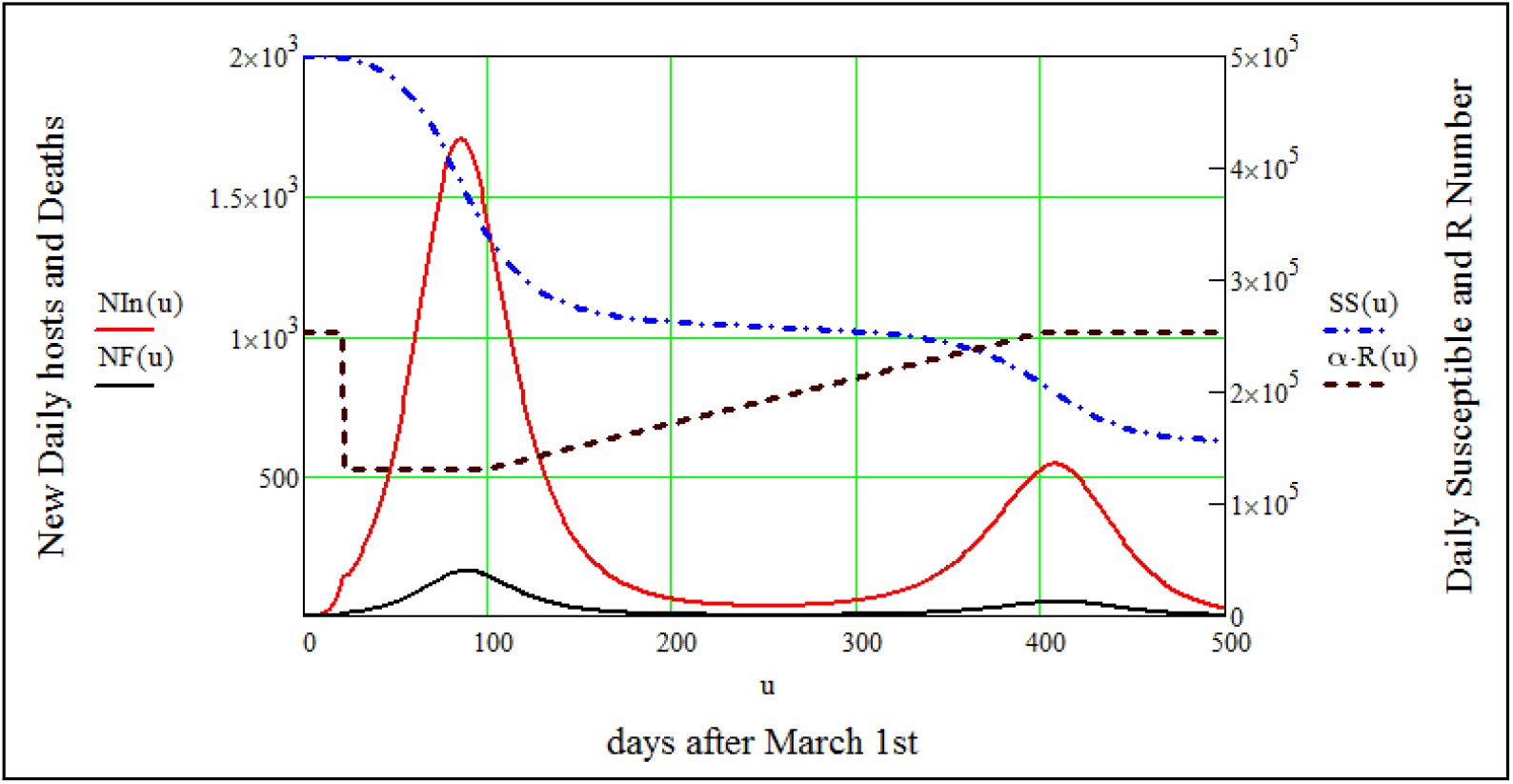
*Result of the MP release over a longer period of time. Simulation by a progressive SD release, starting by the end of the third month, and ending ten months later, as shown by R(t)*.

### Herd immunity analysis

Acquired immunity is conquered at the level of the individual, either through natural infection with a pathogen or through immunization with a vaccine. Herd immunity stems from the effects on individual immunity scaled to the level of the whole population. It is referred to the indirect protection from infection conferred to susceptible individuals when a sufficiently large proportion of immune individuals exist in a population. Depending on the prevalence of existing immunity to a pathogen in a population, an infected individual propagates the disease through susceptible hosts, following effective exposure to infected individuals as described by the SEIR model. However, if a percentage of the population has acquired some immunity level, the likelihood of an effective contact between infected and susceptible hosts is reduced, and the infection will not transmitted by this path. The threshold proportion of susceptible persons required for transmission is known as the, or critical proportion P_c_ [15].

A relevant measure to evaluate the social cost of achieving global SARS-CoV2 herd immunity is the use of the Causality Rate (CR), defined as the proportion of deaths caused by a certain disease among all infected individuals. Now days in Brazil many Covid-19 cases are not reported, especially among asymptomatic hosts or individuals with mild symptoms, the CR will inherently be lower due to sub-notifications. It is important to remember that was established 50% asymptomatic hosts to the SEIR Model in this study. Massive serological testing will be required to better determine how many individuals have been infected, how many are immune, and how far we are from reaching the herd immunity threshold.

Within all those limitations, we can estimate a value for the herd immunity threshold P_c_. Under the deterministic SEIR model, P_c_ = 1 − 1/R_eff_, i.e., herd immunity threshold depends on a single parameter, the effective basic reproduction number R_eff_ [14]. Where *R_eff_= (1 + r/σ_o_).(1* + *r/γ_o_)*, r is the rate of the initial exponential growth, σ_o_ the exposed rate and γ_o_ the rate to be removed from the symptomatic infected group [14]. Since the onset of SARS-CoV-2 spread, studies have estimated the value of R_eff_ in the range of 1.1 < R_eff_ < 6.6 [16]. From the previous values determined for r, σ_o_, and γ_o_, we obtained R_eff_ = 3.0 ± 0.3, and P_c_ = 0.67 ± 0.03. Again, in this study, R_eff_ is restricted to symptomatic hosts only, i.e., 50% of the exposed ones [17]. As a result, the herd immunity threshold will be ξ_a_.P_c_ = 0.34 ± 0.03, and at least 35% of the population considered here remains to be immunized. As commented before the number of COVID-19 notifications do not include the asymptomatic hosts or individuals with mild symptoms. As a result the number N of susceptible was fold by a factor of three, representing 12.3% of the Sao Paulo population. An extra fraction of 35% or, in the total, over 2 Million of exposed individuals to SARS-CoV-2 are required to cross herd immunity threshold at the city of S. Paulo.

Finally, given that the CR of COVID-19 estimated here is 0.23%, and preserving a factor of three fold in the number *N* of susceptible hosts, 44,000 is estimated as the number of people who could potentially die from COVID-19, whilst the population naturally reaches herd immunity. This number is difficult to be accepted, so, before a new vaccine becomes available reinforcements of mitigation policy become imperative.

## Data Availability

I declare that all data referred to in the manuscript, and note links below were provided by the Brazilian Federal Government (Health Minister), and the State S. Paulo Government as coated in references.

## Notes

### Competing Interest Statement

The authors have declared no competing interest.

### Clinical Trial

I understand that all clinical trials and any other prospective interventional studies must be registered with an ICMJE-approved registry, such as ClinicalTrials.gov. I confirm that any such study reported in the manuscript has been registered and the trial registration ID is provided

### Funding Statement

no external funding was received.

